# Endometrial microbiota composition is associated with reproductive outcome in infertile patients

**DOI:** 10.1101/2021.02.05.21251207

**Authors:** Inmaculada Moreno, Iolanda Garcia-Grau, David Perez-Villaroya, Marta Gonzalez-Monfort, Mustafa Bahçeci, Marcelo J. Barrionuevo, Sagiri Taguchi, Elena Puente, Michael Dimattina, Mei Wei Lim, Georgina Meneghini, Mira Aubuchon, Mark Leondires, Alexandra Izquierdo, Martina Perez-Olgiati, Alejandro Chavez, Ken Seethram, Davide Bau, Carlos Gomez, Diana Valbuena, Felipe Vilella, Carlos Simon

## Abstract

**Background:** Previous evidence indicates associations between the female reproductive tract microbiome composition and reproductive outcome in infertile patients undergoing assisted reproduction. We aimed to determine whether the endometrial microbiota composition is associated with reproductive outcomes of live birth, biochemical pregnancy, clinical miscarriage, or no pregnancy.

**Methods:** Here we present a multicentre prospective observational study using 16S rRNA gene sequencing to analyse endometrial fluid and biopsy samples before embryo transfer in a cohort of 342 infertile patients asymptomatic for infection undergoing assisted reproductive treatments.

**Results:** A dysbiotic endometrial microbiota profile composed of *Atopobium, Bifidobacterium, Chryseobacterium, Gardnerella, Haemophilus, Klebsiella, Neisseria, Staphylococcus* and *Streptococcus* was associated with unsuccessful outcomes. In contrast, *Lactobacillus* was consistently enriched in patients with live birth outcomes.

**Conclusions:** Our findings indicate that endometrial microbiota composition before embryo transfer is a useful biomarker to predict reproductive outcome, offering an opportunity to further improve diagnosis and treatment strategies.

## INTRODUCTION

Humans have co-evolved as holobionts with microbial companions including bacteria, viruses, fungi, yeast and archaea (Simon et al. 2019). These microbes and their genetic information (the microbiome) are now well-characterised; the Human Microbiome Project has revealed that approximately 9% of the total human microbiome is found in the female reproductive tract (Peterson et al. 2009). Historically, all microbes were believed to inhabit only the lower portion of the tract; the cervix was considered a perfect barrier between the vagina and the upper genital tract that maintained sterility of the uterine cavity (Tissier 1900). However, accumulating evidence demonstrates that this tract is an open system with a microbiota continuum that gradually changes from the outer to the inner organs, with decreasing bacterial abundance and increasing bacterial diversity from the vagina to the ovaries (Walther-António et al. 2016; Miles, Hardy, and Merrell 2017; C. Chen et al. 2017; Koedooder, Mackens, et al. 2019; Peric et al. 2019). There are reportedly 10^2^–10^4^ fewer bacteria in the uterine cavity than in the vaginal microbiota (Mitchell et al. 2015; C. Chen et al. 2017). Thus, the uterine cavity contains a low-abundance bacterial community, also known as a low-biomass microbiota.

The presence of different microorganisms in the female and male reproductive tracts might influence reproductive function (Franasiak and Scott 2015; Koedooder, Mackens, et al. 2019; Ravel, Moreno, and Simón 2020). Recent studies indicate that the chance of becoming pregnant before the start of an in vitro fertilisation (IVF) treatment is stratified based on the vaginal microbiota composition, as women with a low percentage of *Lactobacillus* in their vaginal sample are less likely to have a successful embryo implantation (Koedooder, Singer, et al. 2019). Analyses by 16S ribosomal RNA (16S rRNA) gene sequencing of endometrial samples suggest that this microbiome impacts reproductive outcomes in infertile patients. Having a *Lactobacillus*-dominated microbiota, defined as ≥90% or ≥80% *Lactobacillus* spp., is associated with significantly increased implantation, pregnancy, ongoing pregnancy, and live birth rates (Moreno et al. 2016; Kyono et al. 2019). Lately, 16S rRNA and whole metagenomics sequencing have been used to investigate the endometrial microbiome (EM) in cases of spontaneous clinical miscarriage (Moreno et al. 2020), recurrent miscarriage (Garcia-Grau et al. 2019), and at 4 weeks gestation in a pregnancy resulting in a live birth (Moreno et al. 2020). Interestingly, the EM during early successful pregnancy exhibited no bacterial diversity and higher *Lactobacillus* abundance (Moreno et al. 2020).

In this prospective multicentre observational study, we investigated the EM composition in 342 infertile patients undergoing assisted reproductive technology (ART) in 13 different centres on 3 continents by analysing endometrial fluid (EF) and endometrial biopsy (EB) samples using 16S rRNA sequencing. The objective of the study was to determine whether EM composition is associated with reproductive outcomes: live birth (LB), biochemical pregnancy (BP), clinical miscarriage (CM), or no pregnancy (NP). *Lactobacillus* was consistently more abundant than dysbiotic or pathogenic bacteria in women that achieved a pregnancy with a LB. *Lactobacillus* spp. depletion and increased abundance of specific taxa including *Gardnerella, Haemophilus, Klebsiella, Neisseria, Staphylococcus, Streptococcus, Atopobium, Bifidobacterium*, and *Chryseobacterium* were associated with NP or CM. The microbiota composition in EF did not fully reflect that in EB, although their association with clinical outcome was consistent.

## METHODS

### Study design and sample size calculation

This was a multicentre prospective observational study analysing the EM of infertile patients with maternal age ≤40 years undergoing IVF or ≤50 years undergoing ovum donation. The EM was analysed in the same endometrial sample in which endometrial receptivity analysis (ERA) was conducted in a cycle prior to embryo transfer of frozen blastocyst stage embryos (day 5/6). Patients were all assessed in a hormone replacement therapy cycle after 120 h of progesterone administration. The EF sample was aspirated before EB collection for EM analysis by 16S rRNA sequencing. The EB was divided into two pieces for the study of endometrial receptivity and EM analysis. Patients were treated following the standard protocols in each clinic and the embryo was transferred on the day recommended by the ERA test result using the same hormonal protocol as for the diagnostic cycle.

The main objectives of this prospective study were to validate our pilot study of the endometrial microbiome and its impact on assisted reproduction in a larger number of patients (Moreno et al. 2016), and to study the topologic effect of the endometrial microbiome by comparing the microbial populations found in EF versus EB. To achieve these objectives, we relied on the formula described for estimating the difference between two proportions (hypothesis of contrasts in unilateral sense), with implantation, pregnancy and ongoing pregnancy rates expected to be about 20% higher in patients with a normal microbiome than in patients with an altered microbiome (95% CI and 80% of statistical power). To detect this difference, we calculated that we would need to assess 146 patients. To validate and compare the two endometrial sample types, we used the endometrial tissue as the gold standard, and the most known and standardised test (ERA). We aimed to assess at least 100 non-receptive patients and 234 receptive patients to obtain a sensitivity and specificity of 90% (estimated using 10 cases in each marginal box of a 2×2^4^ table). Because this last calculation requires the greatest number of patients, we added 30% for possible losses, bringing the total target number of patients to 434.

### Study population — inclusion and exclusion criteria

From August 2017 to February 2019, 452 participants were recruited from 13 reproductive clinics in Europe, America, and Asia. As this was a competitive study, each centre recruited patients and sent their samples to Igenomix Foundation until the target sample size of the study was reached. Inclusion criteria were patients undergoing IVF aged ≤40 years or patients undertaking ovum donations aged ≤50 years; BMI of 18.5–30 kg/m^2^ (both inclusive); negative serological tests for human immunodeficiency virus, Hepatitis B and C viruses, and syphilis; regular menstrual formula (3–4 / 26–35 days); and >2 million sperm/mL. Exclusion criteria included carriers of intrauterine devices or patients who took antibiotics in the last 3 months before sample collection (except mandatory prophylactic antibiotic treatment before egg retrieval, which occurred one month before sample collection); presence of uncorrected adnexal or uterine pathologies as uterine malformations; patients with severe or uncontrolled bacterial, fungal or viral infections, or any illness or medical condition that risks the patient’s safety.

### Sample collection

EF and EB were collected following the previously described procedures (Vilella et al. 2013; Simón et al. 2020). Briefly, with the patient in the lithotomy position, the vagina and cervix were cleaned. After introduction of the disinfected speculum, a sterile and flexible catheter (the same one used for embryo transfer) (Gynétics, Lommel, Belgium) was introduced into the uterine cavity to aspirate EF, obtaining a volume between 20 and 80 μL. To prevent contamination, any contact with vaginal walls was avoided and suction was stopped at the entrance of the internal cervical os during catheter removal. Then, an EB sample (about 50– 70 mg tissue) was obtained with a cannula of Cornier (CCD Laboratories; Paris, France) by scraping the endometrium. Once the EF and EB samples were obtained, they were transferred to cryotubes containing 50 μL and 1.5 mL of RNAlater solution (Qiagen, Hilden, Germany), respectively. Both samples were stored at 4°C for 4 hours, subsequently sent to Igenomix at room temperature and stored at −80°C until use.

### Endometrial receptivity diagnosis

The ERA test (https://www.igenomix.com/genetic-solutions/era-endometrial-receptivity-analysis/) was performed on the human RNA in each obtained EB sample. This test is used to assess the personalised window of implantation of a given patient and determine the optimal time frame for embryo transfer (Díaz-Gimeno et al. 2011; Ruiz-Alonso et al. 2013; Simón et al. 2020).

### DNA isolation

Total DNA was isolated from EF samples by performing a pre-digestion step at 37°C for 30 minutes with 25 ug/μL lysozyme, 0.12 U/μL lysostaphin, 0.4 U/μL mutanolysin and 1.8% Triton X-100 to degrade the bacterial cell walls. DNA was then extracted with the QIAamp DNA Blood Mini kit (Qiagen) following the manufacturer’s instructions. Finally, the DNA was eluted with 35 μL of nuclease free water and quantified using a photometric technology (Nanodrop, Waltham, MA, USA).

Total DNA was isolated from the EB samples by performing a pre-digestion step for difficult-to-lyse bacteria. For this digestion, 25 mg of tissue was cut into small pieces and treated with proteinase K at 56°C for 3 hours under agitation. Samples were then mixed with ATL buffer and disrupted mechanically in a TissueLyser LT for 5 minutes at 50 Hz using stainless-steel beads (all acquired from Qiagen). After these pretreatments, bacterial nucleic acids were purified using the DNA tissue program V7-200-LC of the QIAsymphony (Qiagen) following the manufacturer’s instructions. Finally, the DNA was eluted with 50 μL of nuclease free water and quantified using MultiskanGO (Thermo Scientific, Waltham, MA, USA).

### 16S ribosomal RNA gene sequencing

The EM profiles were obtained by next-generation sequencing using the Ion 16S metagenomics kit (Thermo Fisher Scientific), which selectively amplifies 7 of the 9 hypervariable regions of the bacterial gene encoding the 16S ribosomal subunit (V2-4-8 and V3-6, 7-9). After amplification of the hypervariable regions with 10 μL of sample (per set of primers) and 30 PCR cycles, the library was prepared starting from 50 ng of the pooled short amplicons using the Ion Plus Fragment Library kit and Ion Xpress Barcode Adaptors following the manufacturer’s instructions. The library concentration was adjusted using the Ion Universal Library Quantitation Kit and QuantStudio 5 Real-Time PCR System. The diluted individual libraries were then pooled for amplification by emersion PCR in the Ion OneTouch 2 System (10 pM) or Ion Chef System (30 pM). Finally, libraries were sequenced on the Ion S5 XL system using the Ion 530 Chip (all acquired from Thermo Fisher Scientific).

To detect eventual contamination, each sequencing run included between two and four blank samples as well as negative and positive PCR controls. The blank samples consisted of an aliquot of the sample preservation buffer (RNA later; Qiagen), while positive and negative PCR controls were pure microbial DNA from *E. coli* (3 ng) and nuclease-free water, respectively (Thermo Fisher Scientific).

### Sequencing-based microbiome analysis

The 16S rRNA sequencing results were analysed using the QIIME 2.0 package (https://qiime2.org) and RDP classifier 2.2 for taxonomic assignment along with the greengenes database version 13.8 (http://greengenes.second.genome.com), an update of version 13.5 released to address missing genus and species names. For all processed samples, the RDP classifier was run with the default minimum confidence estimate of 0.5 to record an assignment (i.e., an operational taxonomic unit [OTU]). The results were generated using the default parameters in the original QIIME 2 (Bolyen et al. 2019) and RDP (Wang et al. 2007) methods. Identification and removal of contaminant sequences was conducted using the decontam R Bioconductor package (Davis et al. 2018), using bacterial prevalence in blank samples for statistical identification of contaminant taxa. Data were transformed to log-ratios using the clr transformation (Aitchison 1986) making them symmetric and linearly related, and thus potentially avoiding spurious correlations and sub-compositional incoherencies (Calle 2019; Gloor et al. 2017; Pawlowsky-Glahn, Egozcue, and Tolosana-Delgado 2015).

Taking into account quality parameters such as the percentage of empty reads, the dispersion index, and the ratio between filtered and mapped reads, samples were classified as detectable or not detectable biomass based on the filtered versus mapped reads ratio threshold of 0.65 in EF and 0.7 in EB samples. For further analysis, only the detectable samples (i.e. samples with filtered versus mapped reads ratios greater than 0.65 and 0.7 in EF and EB, respectively) were included. In each dataset of endometrial 16S rRNA profiles, the abundance of each taxa was analysed, and low-abundance species were removed from further consideration. We retained taxa that either (1) exhibited an abundance of at least 1% in 5% of the samples or (2) exhibited an abundance of at least 0.1% in at least 15% of samples (Fettweis et al. 2019). Taxa that failed to meet both criteria were removed. Genera not colonising humans or associated with kitome contaminants were also removed from the analysis. Specifically, taxa consistently reported to be contaminants in more than 5 of the 11 published references were excluded (Tanner et al. 1998; Grahn et al. 2003; Barton et al. 2006; Laurence, Hatzis, and Brash 2014; Lauder et al. 2016; Glassing et al. 2016; Stinson, Keelan, and Payne 2019; Weyrich et al. 2019; Kyono et al. 2018; Hashimoto and Kyono 2019; Kitaya et al. 2019). After applying these criteria, 15 and 19 OTUs were evaluated in EF and EB microbiota respectively, with 12 OTUs shared between sample types (Supplementary Table 4).

### Correlation network analysis

Pearson matrices for network analysis were generated using the Scipy Stats package (version 1.4.1) and visualised with Networkx (version 2.4), considering taxa with significant Pearson correlation coefficient (Li et al. 2008). To account for both the correlation and significance, each edge was assigned a weight w computed as: abs(corr) – pval, where abs(corr) was the absolute correlation value between each pair of taxa and pval was the corresponding p-value. Negative correlations were represented as red edges. Network analysis was performed on each data type (i.e., EF and EB) and for each outcome (i.e., LB, NP, BP, and CM).

### Taxa reference ranges

The log-ratio-transformed 16S rRNA profiles from LB samples were used to define the reference range for each taxon. A confidence interval of 95% was calculated to establish reference ranges for the Student’s t-distribution taxa distribution model (Almonacid et al. 2017). The Scipy Stats package (version 1.4.1) was used for calculations (Virtanen et al. 2020).

Taxa contributions to different outcomes were statistically inferred using a two-sided Mann-Whitney U-test. The difference between samples outside the range and upper/lower bound reference values was used to assess the role of the taxa in each outcome. Taxa were reported when a significant difference was observed between upper and lower difference distributions. The Scipy Stats package (version 1.4.1) (Virtanen et al. 2020) was used for statistical methods and Plotly was used to visualise significant taxa distributions (Inc. 2015).

### Bayesian inference for reproductive outcome differences

Differences between *Lactobacillus* and other reproductive tract taxa were modelled using Bayesian inference following a normal distribution with mean *µ*_*outcome*_ and dispersion *σ*_*outcome*_ for each reproductive outcome,

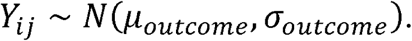

Priors selected to estimate parameters were normal distribution for *µ*_*outcome*_ centred on the mean difference between *Lactobacillus* and other taxa in the dataset and a dispersion of 10 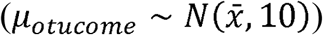 and, for *σ*_*outcome*_, vague continuous distribution from 0 to 10 (*σ*_*outcome*_ ∼ *Uniform*(0,10). Markov chain Monte Carlo sampling for posterior distribution parameter inference and predictive probability analysis were implemented using the PyMC3 Python library (Salvatier, Wiecki, and Fonnesbeck 2016).

### Statistical analysis

All data were analysed on a per-protocol basis. One-way analysis of variance was used to compare non-categorical variables among groups by reproductive outcomes. Mean differences and standard deviation or median and interquartile ranges were used when the variables were not homogeneous, as well as the mean differences with 95% CI values. Categorical variables were described by counts (n) and percentages (%), and the Chi-square test and two-sided Fisher’s exact test were used to compare groups by reproductive outcomes with respect to percentages. Multiple-comparison post-hoc correction (Bonferroni) was applied for all pairwise comparisons.

Multinomial logistic regression analysis was conducted to control possible confounding factors, effect modifiers, to demonstrate the homogeneity of the key variables and the absence of bias towards our final endpoint. A multinomial endpoint was included as a dependent variable (LB/NP/BP/CM) and predictors shown in Supplementary Table 1 and 2. P < 0.05 was considered to be statistically significant. All analyses were conducted using SPSS 25 software (IBM, MD, USA) and R version 3.6.3 (The CRAN project).

## RESULTS

### Patient cohort, characteristics, and outcomes

A total of 452 patients with infertility undergoing IVF were assessed for eligibility in 13 reproductive clinics in Europe, America, and Asia between August 2017 and February 2019 (Figure 1). Forty-four patients were excluded from the study because they did not meet the inclusion criteria (n=42) or declined to participate (n=2). The remaining 408 women were recruited and their EB and EF microbiota composition was assessed by 16S rRNA sequencing. However, 66 patients were lost to follow-up. Of the 342 remaining patients, 198 (57.9%) became pregnant [141 (41.2%) had a LB, 27 (7.9%) had a BP and 28 (8.2%) a CM], while 144 (42.1%) did not become pregnant (Supplementary Table 1). Moreover, 2 patients experienced an ectopic pregnancy, but their results were not considered for further comparisons due to the small sample size (Figure 1). Analysed patients had a mean age of 36 years (range 21–49), and a mean body mass index (BMI) of 23.3 (range 18.5–30.0). The ethnic distribution was Caucasian (57.3%), East Asian (14.0%), Hispanic (11.4%), and others (17.3%). The indications for IVF were advanced maternal age, male factor infertility, unexplained infertility, and ovarian pathology. The assessed clinical and embryological variables displayed homogeneity and no bias toward the clinical outcome categories was observed (Supplementary Table 2).

**Figure 1.**
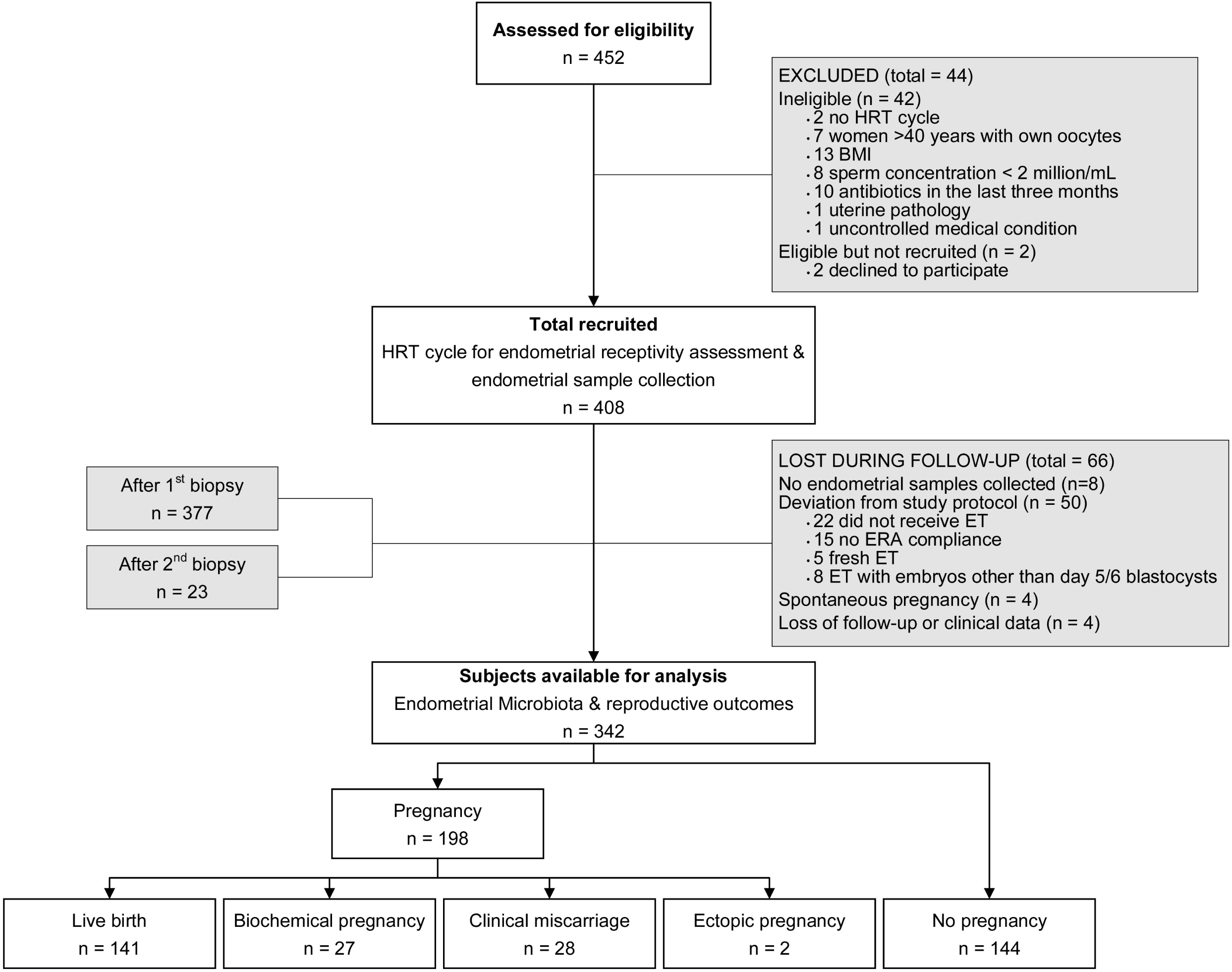
Flowchart of the study population. Of all patients assessed for eligibility (n = 452), 44 were excluded from the analysis and 66 were lost to follow-up. Thus, 342 patients were ultimately included in our assessment of the impact of the endometrial microbiome on pregnancy outcomes. Abbreviations: BMI: body mass index; ERA: endometrial receptivity analysis; ET: embryo transfer; HRT: hormonal replacement therapy.

### Endometrial microbiota composition in EF and EB

The EM was profiled in EF from 336 patients and EB for 296 patients, with paired EF–EB results in 290 (84.8%) participants. The mean total sequencing reads per sample was 302,299 (range, 110,050–394,659) in EF samples, and 335,659 (range, 237,889–430,675) in EB samples, with an average of 89,883 (range, 27,960–137,956) and 103,539 filtered reads (range, 61,650–162,653), respectively (Supplementary Table 3).

Because the endometrium presents a low-abundance microbiota, stringent analysis was performed to ensure that contaminating reads did not interfere with downstream analysis. Samples were classified as detectable and not-detectable by comparing them to blank samples included in each run and assessing certain quality parameters (see criteria in Materials and Methods). Despite starting with equivalent amounts of extracted DNA, detectable samples showed a different clustering behaviour as compared with not-detectable/low-biomass samples (with a higher 16S amplicon concentration), which clustered together with blank controls (Figure 2). After applying these criteria, 208 EF samples and 190 EB samples were classified as detectable and included in the analysis (Figure 2).

**Figure 2.**
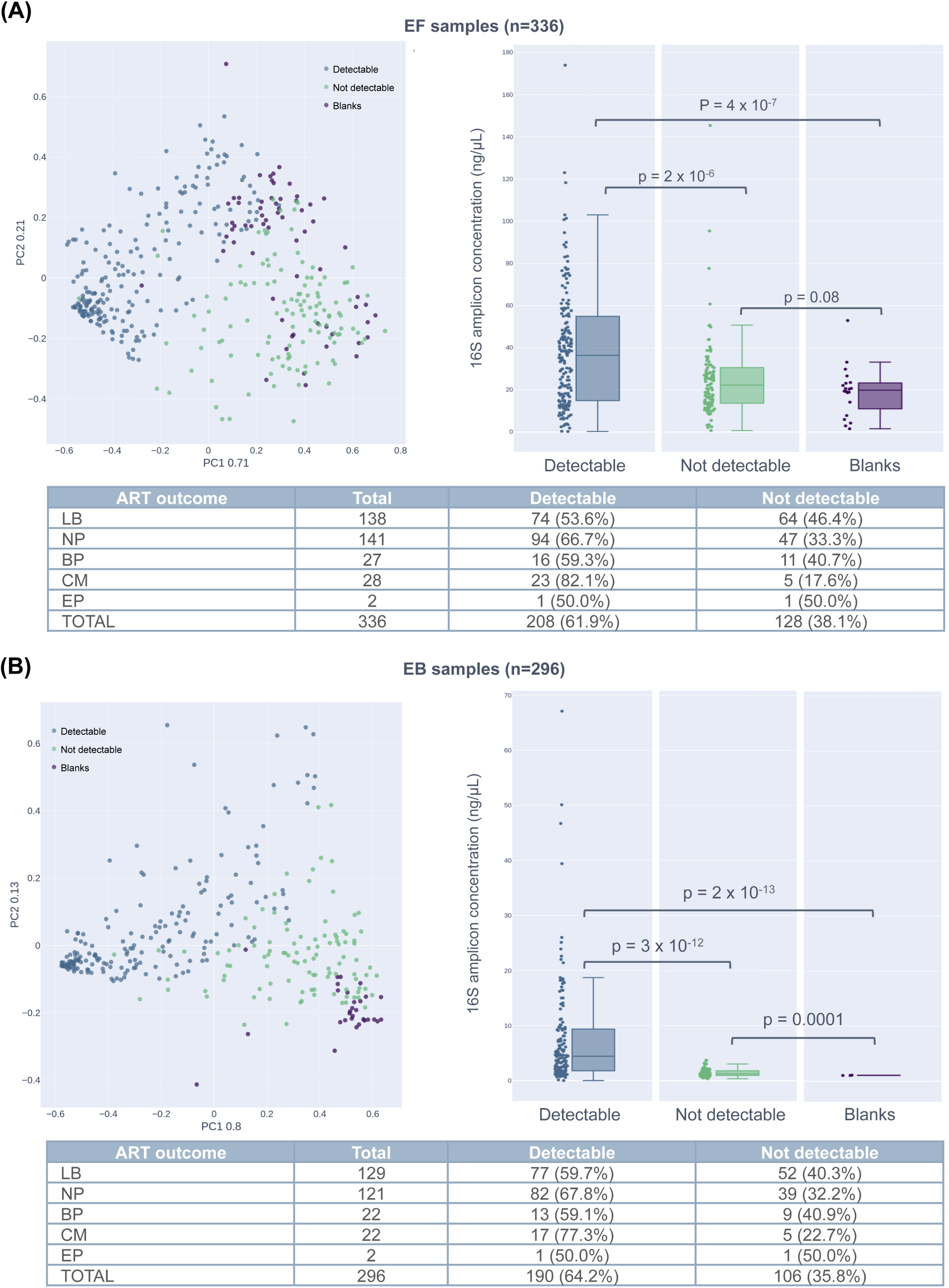
Distribution of sequencing data. PCA showing the clustering of the (A) endometrial fluid samples (n=336) and (B) endometrial biopsy samples (n=296) and their corresponding blank controls, based on quality parameters such as percentage of empty reads, dispersion index for each sample and the ratio between the filtered and mapped reads. Samples are coloured using a filtered/mapped reads threshold of 0.65 for EF and 0.7 for EB. Abbreviations: BP, biochemical pregnancy; EP, ectopic pregnancy; LB, live birth; CM, clinical miscarriage; NP, no pregnancy.

*Lactobacillus* was the major genus in both EF and EB samples. Bacterial genera such as *Anaerococcus, Atopobium, Bifidobacterium, Corynebacterium, Gardnerella, Haemophilus, Microbacterium, Prevotella, Propionibacterium, Staphylococcus*, and *Streptococcus* were also commonly identified in both sample types (Supplementary Figure 1). *Streptomyces, Clostridium*, and *Chryseobacterium* were detected in EF but not EB, whereas *Cupriavidus, Escherichia, Klebsiella, Bacillus, Finegoldia, Micrococcus*, and *Tepidimonas* were detected in EB but not EF (Supplementary Figure 1).

The co-occurring EM bacterial networks showed several differences between the sample types: (i) the EF microbiota had two linked communities, while the EB microbiota had four linked communities and two isolated nodes; (ii) the EF microbiota network was more strongly connected than the EB community; and (iii) *Lactobacillus* had positive and negative connected neighbours in the EF microbiota, but only negative relations in the EB microbiota (Figure 3). In the EB microbiota, *Lactobacillus* was negatively correlated with pathogenic bacteria *Gardnerella, Bifidobacterium*, and *Atopobium*, whereas in EF, *Lactobacillus* was negatively correlated with *Gardnerella, Bifidobacterium, Atopobium, Staphylococcus, Streptococcus*, and *Chryseobacterium*, and positively correlated to commensal bacteria (*Clostridium* and *Streptomyces*).

**Figure 3.**
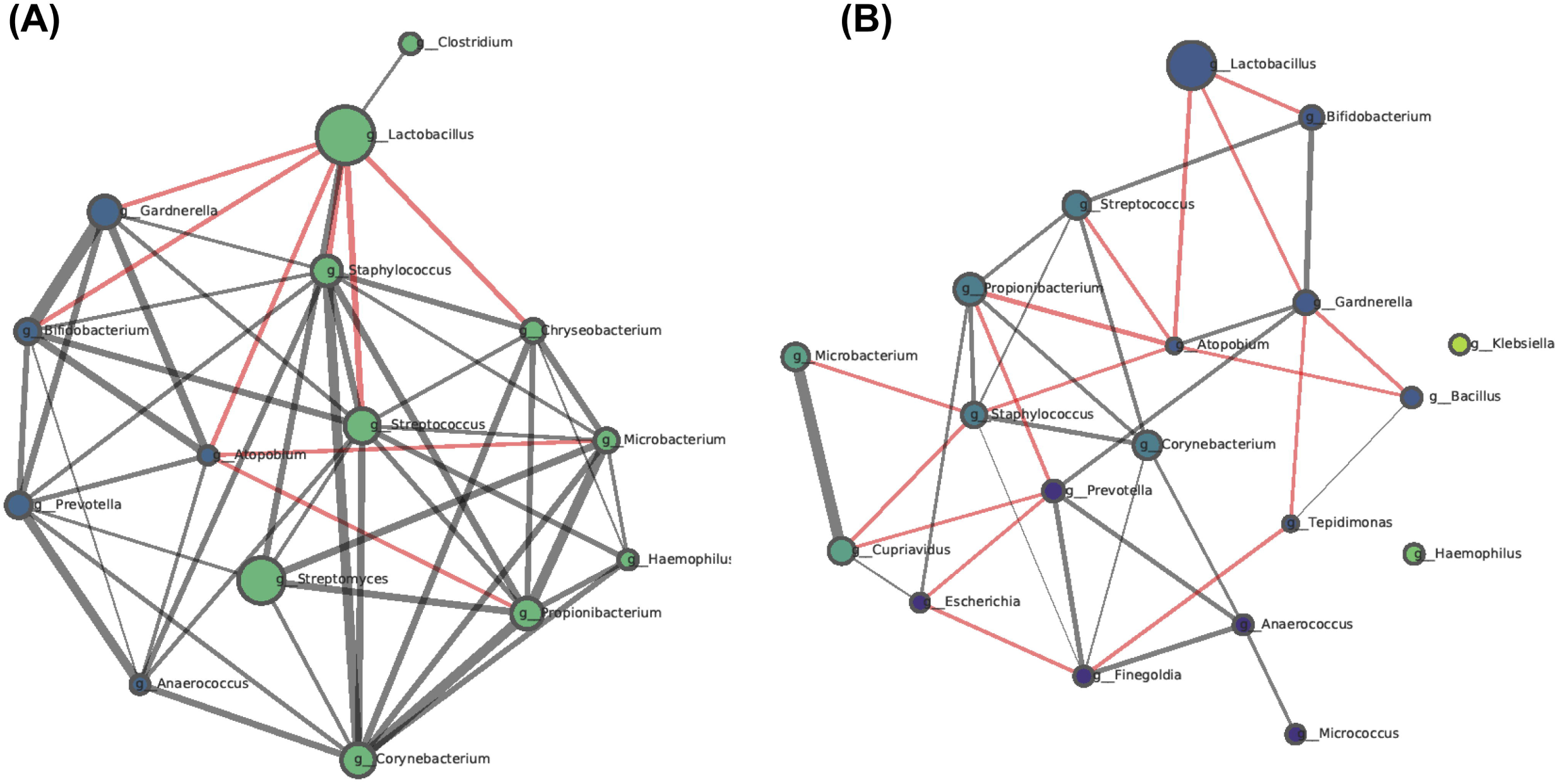
Co-occurrence bacterial networks in endometrial fluid (A) and endometrial biopsy (B) samples. Each network was created by computing the co-occurring bacteria with significant Pearson correlation coefficients. Samples from all reproductive outcomes are represented. Node properties: (i) circle size, proportional to the normalised and standardised bacterial relative abundances; (ii) colour, communities as retrieved by the Louvain algorithm. Edge properties: (i) thickness, proportional to p-value of Pearson correlation coefficient, from the most significant (thicker) to the less significant (thinner); (ii) colour, red for negative, and grey for positive Pearson correlation coefficients.

### Endometrial microbiota composition and reproductive outcome

To analyse the association between EM composition in EF and EB and reproductive outcome, we built microbiota networks for each reproductive outcome. We found that the LB category was denser and had a higher node degree distribution than co-occurrence networks of unsuccessful reproductive outcomes. Additionally, we found potential interactions that only occurred in patients with LB, reflecting the relevance of these relationships to successful pregnancy and how their disruption may lead to ecosystem instability. We also noted that in the EF microbiota of patients who had a LB, *Lactobacillus* was negatively related to dysbiotic bacteria such as *Chryseobacterium, Staphylococcus* and *Haemophilus*, and positively correlated to *Streptomyces*, which in turn was part of a dense community mainly composed of commensal bacteria such as *Corynebacterium, Microbacterium, Propionibacterium*, and *Clostridium*. In patients with NP, we identified a similar behaviour, with *Lactobacillus* negatively correlated with *Gardnerella, Bifidobacterium, Atopobium, Staphylococcus, Streptococcus*, and *Chryseobacterium*, and positively related to *Streptomyces*. Interestingly, in the group of patients with BP and CM, these interactions disappeared, and the resulting networks were disconnected and formed sparse communities (Figure 4A). Finally, the EB microbiota networks were more dispersed than the EF ones, with fewer interactions between *Lactobacillus* and other taxa. Thus, in this case, the eventual beneficial/deleterious connections among taxa were less evident (Figure 4B).

**Figure 4.**
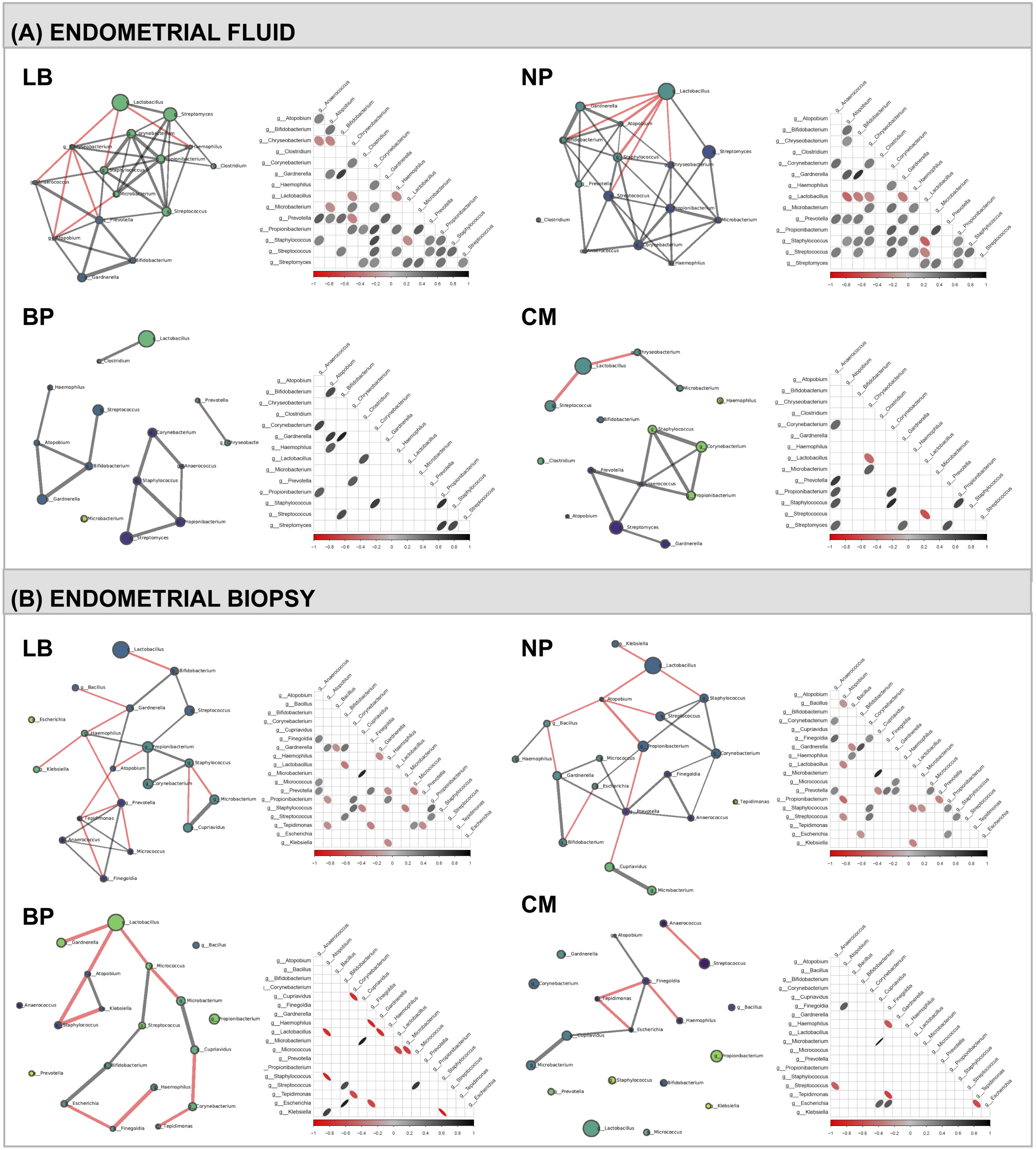
Co-occurrence bacterial networks associated with reproductive outcomes. Co-occurrence bacterial networks in (A) endometrial fluid samples and (B) endometrial biopsy samples for each ART outcome. Each network was created by computing the co-occurring bacterial communities with significant Pearson correlation coefficients. Node properties: (i) circle size, proportional to the normalised and standardised bacterial relative abundances; (ii) colour, communities as retrieved by the Louvain algorithm. Edge properties: (i) thickness, proportional to p-value of the Pearson correlation coefficient, from the most significant (thicker) to the less significant (thinner); (ii) colour, red for negative, and grey for positive Pearson correlation coefficients. For association graphs, the same criteria were applied, with the thickness of the circle and colour intensity being proportional to the corresponding Pearson correlation coefficients. Pairs of bacteria without a circle have no significant Pearson correlation coefficient. BP, biochemical pregnancy; CM, clinical miscarriage; LB, live birth; NP, no pregnancy.

To avoid potential bias when comparing samples analysed in different runs, bacterial profiles were transformed into centred log ratio (clr) data, and the bacterial communities were analysed according to the difference between *Lactobacillus* and other reproductive tract taxa using z-score-normalised values. Using these conditions, higher abundance of *Lactobacillus* was observed in both EF and EB in patients with LB compared to patients with negative reproductive outcomes (Figure 5A). Taxa with a higher average abundance in unsuccessful outcomes than in LB included *Streptococcus, Chryseobacterium, Corynebacterium, Haemophilus, Bifidobacterium, Staphylococcus, Atopobium, Gardnerella*, and *Propionibacterium* in the EF microbiota and *Gardnerella, Klebsiella, Atopobium, Finegoldia, Escherichia, Propionibacterium, Haemophilus, Anaerococcus*, and *Bacillus* in the EB microbiota (Supplementary Figure 2). Predictive probability analysis using a Bayesian inference model showed a different highest posterior density (HPD) interval of the difference “*Lactobacillus* – other taxa” for each ART outcome: LB (−0.12–1.51), NP (−1.05–0.65), BP (−3.45–0.81), and CM (−2.81–0.55). Patients with a LB were more likely to have a higher abundance of *Lactobacillus* (Figure 5B). This increased probability of higher *Lactobacillus* abundance in LB was especially distinct in EF samples, where the HPD for LB samples showed less overlap with unsuccessful outcome intervals.

**Figure 5.**
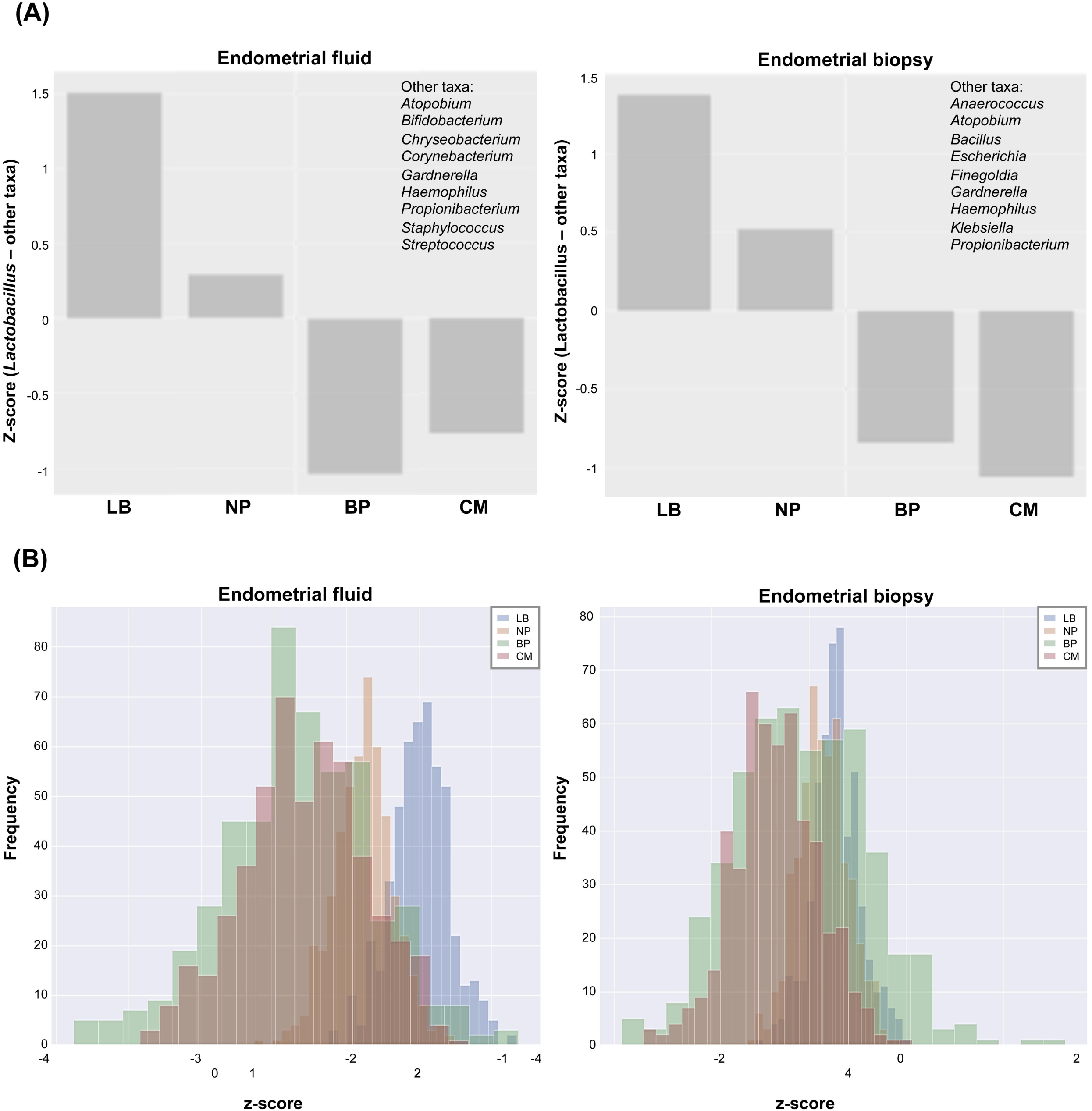
*Lactobacillus* is more abundant than other taxa in reproductive success vs failure. (A) Difference between *Lactobacillus* and other reproductive tract taxa using z-score-normalised values in endometrial fluid (left panel) and endometrial biopsy (right panel) samples. (B) Predictive model showing the probability of each reproductive outcome based on the EM profile. Posterior predictive distribution density plot of z-score differences between *Lactobacillus* and other reproductive tract taxa by reproductive outcome. BP, biochemical pregnancy; CM, clinical miscarriage; LB, live birth; NP, no pregnancy.

Finally, we compared the EM bacterial profiles in patients who achieved a successful pregnancy with LB versus those with unsuccessful outcomes (BP, CM and NP). Our hypothesis was that the microbiota composition in patients with LB is the physiological scenario and does not interfere with functional reproductive potential. Therefore, we evaluated the distance between the abundance of each bacterial taxon and reproductive outcome in the upper and lower 95% confidence intervals established for patients with LB (Figure 6). In patients with NP, the EF taxa with significantly higher abundance exceeding the established upper confidence interval were *Atopobium, Bifidobacterium, Chryseobacterium, Gardnerella*, and *Streptococcus*, and in those with CM, *Haemophilus* and *Staphylococcus* exceeded the physiological levels. By contrast, taxa with a significantly higher distance to the lower established confidence interval were *Lactobacillus* and *Microbacterium* in NP patients, and *Lactobacillus* in patients with CM (Figure 7A). In the EB microbiota of NP patients, *Bifidobacterium, Gardnerella* and *Klebsiella* were significantly more abundant, and the abundance of *Cupriavidus, Finegoldia, Lactobacillus*, and *Tepidomonas* was significantly below the established confidence interval (Figure 7B). The remaining comparisons did not reach statistical significance, possibly due to the small number of patients with BP and CM.

**Figure 6.**
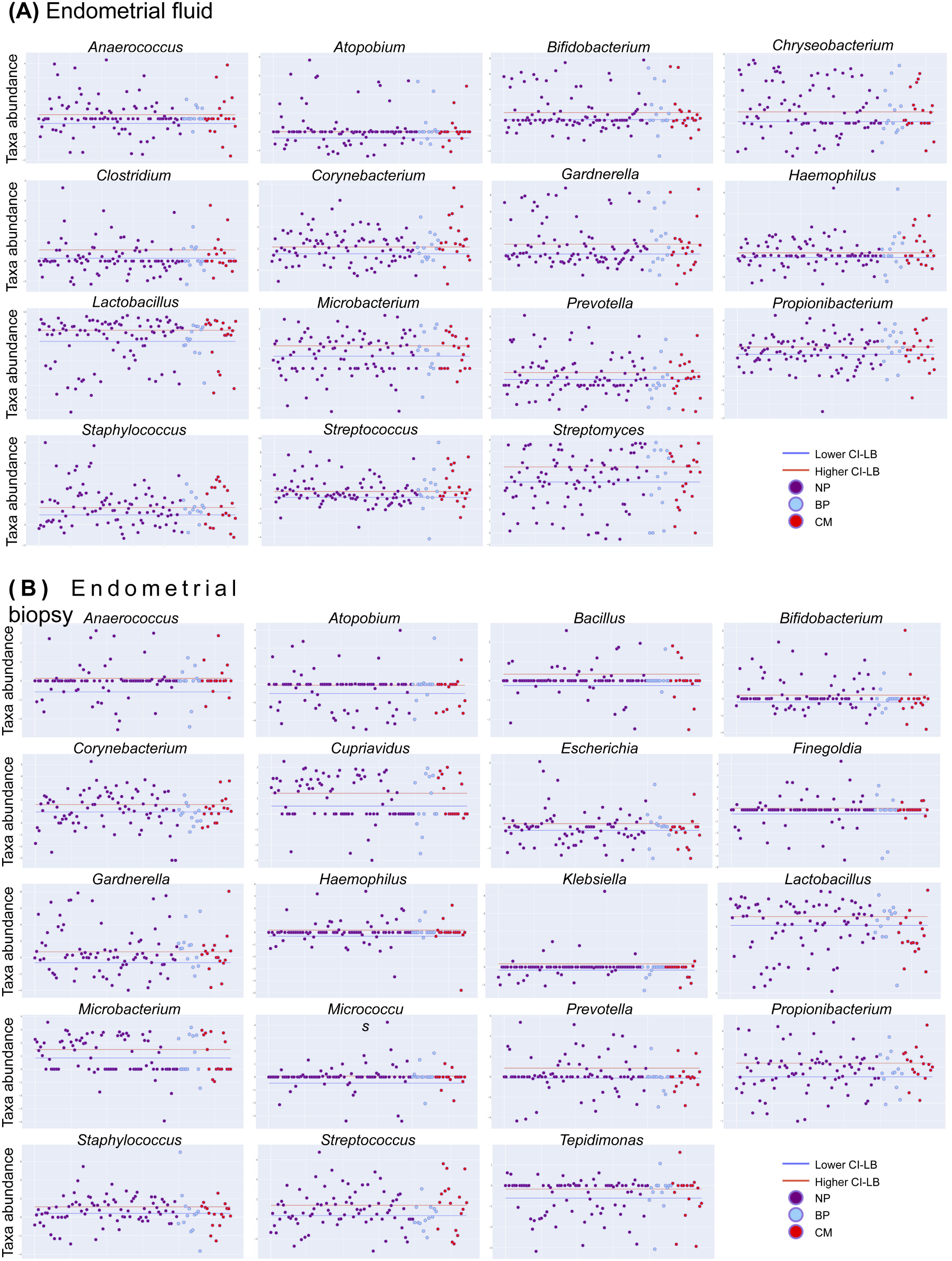
Confidence intervals and reference ranges of bacterial taxa detected in patients with live birth. The endometrial microbiome from patients with a live birth was analysed in (A) endometrial fluid and (B) endometrial biopsy samples to determine the reference ranges for each evaluated taxon. The confidence interval displays the relative abundance for all assessed patients, revealing the healthy ranges of abundance for the taxa in the tested panel. The healthy distribution was used to define the 95% confidence interval (red line) and taxa abundance in patients with poor reproductive outcomes—no pregnancy, biochemical pregnancy, and clinical miscarriage—were comparatively represented. A taxa abundance of 0 was assigned when the bacteria was not detected in a given sample. BP, biochemical pregnancy; CM, clinical miscarriage; LB, live birth; NP, no pregnancy.

**Figure 7.**
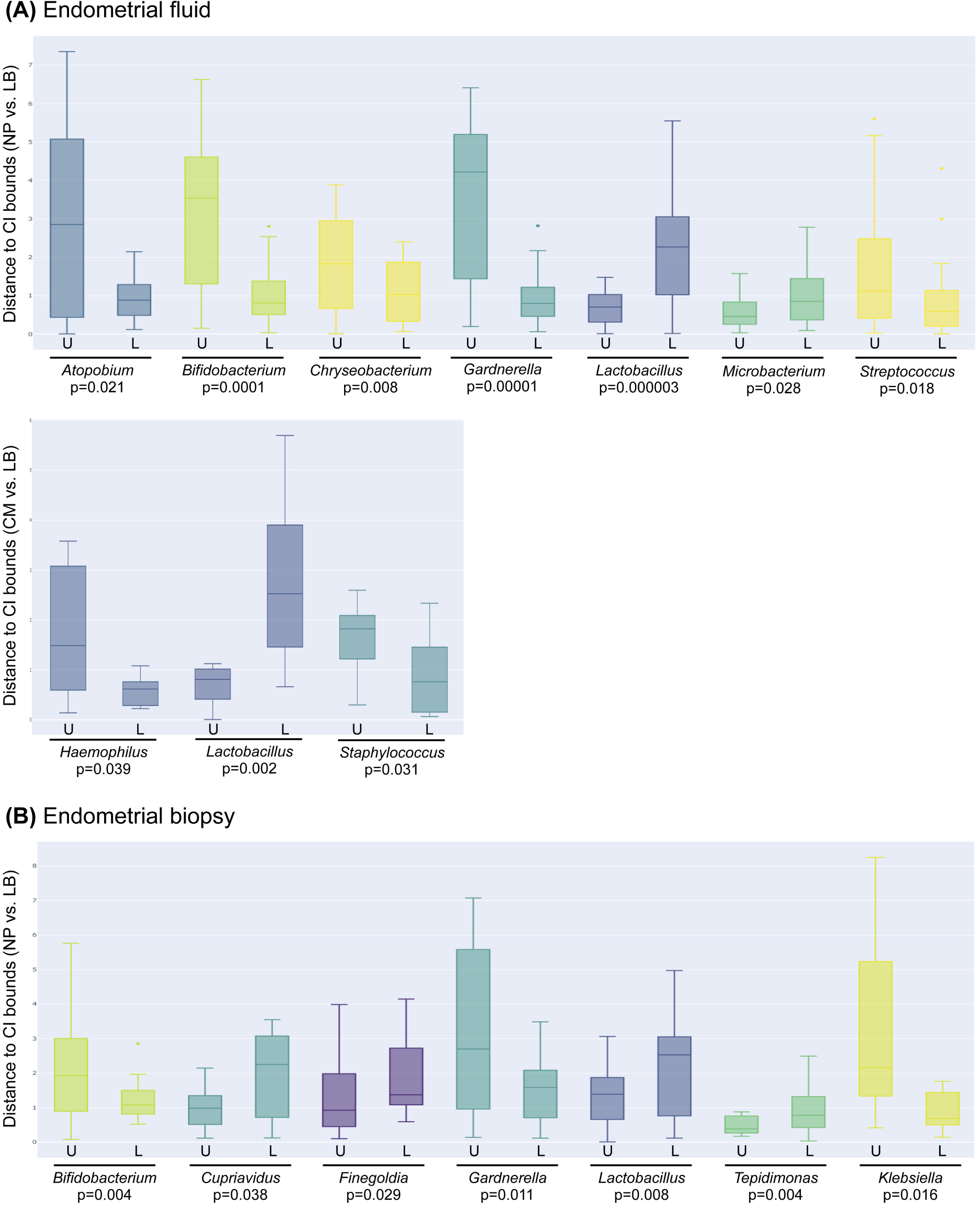
Pathogenic bacterial profiles significantly associated with reproductive outcome. Box plots showing taxa with significant differential abundance in no pregnancy (NP), biochemical pregnancy (BP) and clinical miscarriage (CM) compared to live birth (LB). Differential abundance was calculated using the distance of each value to the upper (U) or lower (L) bounds for the 95% CI in LB (Figure 5) in (A) endometrial fluid and (B) endometrial biopsy samples. Only taxa with significant differential abundance, calculated with a two-sided Mann-Whitney U-test, are represented in the graphs. BP, biochemical pregnancy; LB, live birth; CM, clinical miscarriage; NP, no pregnancy.

### Endometrial microbiota composition in chronic endometritis and reproductive outcome

We also evaluated the abundance of the main pathogenic bacteria reported to cause chronic endometritis (CE): *Enterococcus, Escherichia, Klebsiella, Streptococcus, Staphylococcus, Gardnerella, Mycoplasma, Ureaplasma, Chlamydia*, and *Neisseria*. These bacteria are considered to be a potential cause of infertility as well as obstetric and neonatal complications (Kitaya et al. 2016; Kitaya et al. 2018). We compared the abundance of these bacteria with the confidence interval generated for infertile patients that achieved a LB.

Of the CE pathogens, *Gardnerella, Klebsiella*, and *Streptococcus* were significantly increased in the EF microbiota of NP patients, whereas *Enterococcus* was increased in patients that experienced BPs, and *Klebsiella* and *Staphylococcus* were increased in CM (Figure 8A). In the EB microbiota, *Gardnerella, Neisseria*, and *Klebsiella* were significantly enriched in women with NP compared to those that achieved LB, while *Enterococcus* abundance was below the confidence interval (Figure 8B). In the remaining unsuccessful reproductive categories (BP and CM), no significant taxa were detected. Interestingly, *Gardnerella* and *Klebsiella* were the only common pathogens significantly enriched in both EF and EB from patients with NP.

**Figure 8.**
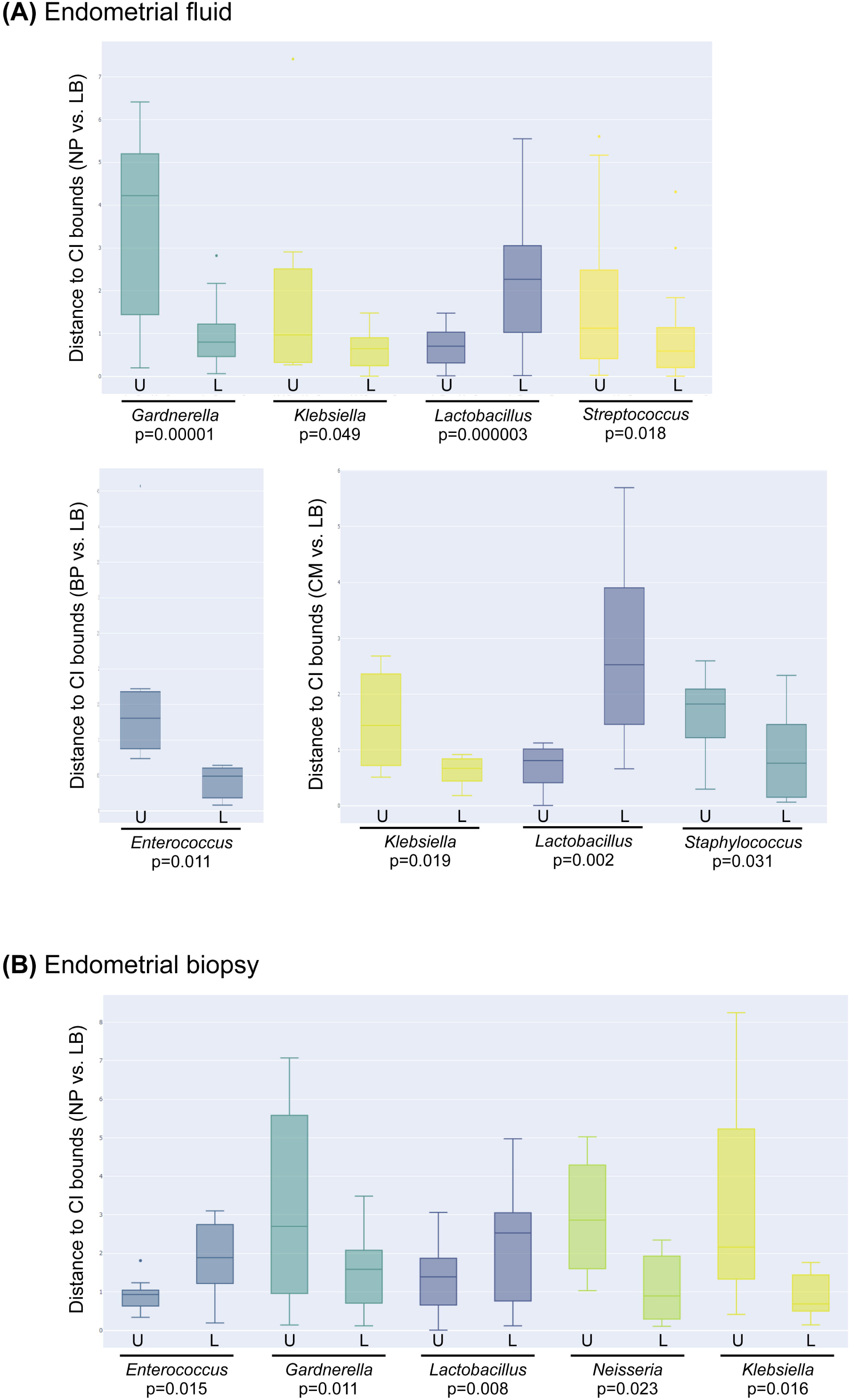
Chronic endometritis profile associated with reproductive outcomes. Box plots showing differential abundance in chronic endometritis-causing bacteria in no pregnancy (NP), biochemical pregnancy (BP) and clinical miscarriage (CM) compared to live birth (LB). Differential abundance was calculated using the distance of each value to the upper (U) or lower (L) bounds for the 95% CI in LB (Figure 5) in (A) endometrial fluid and (B) endometrial biopsy samples. Only taxa with significant differential abundance, calculated with a two-sided Mann-Whitney U-test, are represented in the graphs. BP, biochemical pregnancy; LB, live birth; CM, clinical miscarriage; NP, no pregnancy.

### Endometrial microbiota composition fingerprinting is associated with reproductive outcome

In summary, the pathogenic profile associated with reproductive failure in our cohort of infertile patients consisted of *Atopobium, Bifidobacterium, Chryseobacterium, Gardnerella, Haemophilus, Klebsiella, Neisseria, Staphylococcus*, and *Streptococcus*. In contrast, *Lactobacillus* was consistently enriched in the EM (both in EF and EB) from patients that achieved LB. Therefore, patients with a higher abundance of lactobacilli are more likely to achieve reproductive success. Also, some commensal bacteria such as *Cupriavidus, Finegoldia, Microbacterium* and *Tepidimonas* were positively associated with LB.

## DISCUSSION

Some diseases or organ malfunctions are marked by the presence of potentially pathogenic microbes, whereas others are characterised by depletion of health-associated bacteria. The vaginal microbiota composition is associated with obstetrical outcome (Fettweis et al. 2019; Koedooder, Singer, et al. 2019). However, no consensus has been reached on the profile of endometrial bacterial pathogens and the mechanisms by which they could interfere with embryo implantation (Benner et al. 2018; Baker, Chase, and Herbst-Kralovetz 2018).

In this study, we investigated the relationship between EM composition and the reproductive fate of a cohort of 342 ethnically diverse infertile patients from three different continents. Our results suggest that the composition of the EM at the time of conception is associated with the reproductive outcome. The co-occurrence bacterial association plots built separately for each ART outcome showed that the LB networks were denser and had a higher node degree distribution than the networks of failed outcomes. Moreover, *Lactobacillus* was generally negatively correlated to pathogenic bacteria and positively correlated to commensal bacteria, which may be important for stability in the ecosystem. *Lactobacillus* depletion and the presence of specific pathogenic bacteria such as *Atopobium, Bifidobacterium, Chryseobacterium, Gardnerella, Streptococcus*, and *Klebsiella* in EF, and/or *Bifidobacterium, Gardnerella, Klebsiella*, and *Neisseria* in EB were associated with unsuccessful reproductive outcomes.

*Atopobium vaginae* and *Gardnerella vaginalis* are the major bacterial vaginosis-associated pathogens; they stimulate an innate immune response from vaginal epithelial cells (Libby et al. 2008). However, the deleterious impact of these pathogens is not restricted to the vagina. Women with bacterial vaginosis subjected to laparoscopic salpingectomy or curettage present significantly increased risk of developing endometrial polymicrobial biofilms with *G. vaginalis* and other bacteria (Swidsinski et al. 2013). *Streptococcus agalactiae, Klebsiella pneumoniae, Enterococcus faecalis, Neisseria gonorrhoeae*, and in some studies *G. vaginalis*, are the major pathogens of CE (Kitaya et al. 2018; Cicinelli et al. 2008; Moreno et al. 2018). This condition impairs reproductive outcomes in natural conception and after ART, further contributing to obstetric and neonatal complications (Kitaya et al. 2016). A case series study of three women with recurrent implantation failure and CE showed the elimination of CE-associated pathogens and subsequent restoration of fertility after intrauterine antibiotic administration (Sfakianoudis et al. 2018). Moreover, bacteria such as *S. agalactiae* are known to be one of the leading causes of neonatal infections by vertical transmission (Patras and Nizet 2018). Finally, in the absence of *Lactobacillus, Bifidobacterium* are hypothesised to be able to maintain healthy vaginal balance by the production of lactic acid (Freitas and Hill 2017; Kyono et al. 2019). However, our results show a negative association of *Bifidobacterium* with LB, in agreement with other reports demonstrating *Bifidobacterium* species to be pathogenic in various infectious conditions (Bhaskar, Sistla, and Kumaravel 2017; Pathak, Trilligan, and Rapose 2014; Y. Chen et al. 2019). Hence, the role of members of this genus should be further investigated.

We observed a close relationship between EF and EB microbiota, although there were some differences between the sample types. Specifically, 12 of the 15 most abundant taxa detected in EF overlapped with the taxa identified in the EB samples, but three taxonomies were unique for EF (*Streptomyces, Clostridium*, and *Chryseobacterium*) and seven were detected only in EB (*Cupriavidus, Escherichia, Klebsiella, Bacillus, Finegoldia, Micrococcus*, and *Tepidimonas*). A possible explanation for the difference in bacterial taxa between the sample sources is that bacteria present on the surface of the luminal epithelium may be different than those found deeper, close to the glandular epithelium and stroma. However, these differences may also be due to the different processing and DNA extraction protocols used for the different sample types. The occurrence bacterial networks also revealed differences between the sample types. The EB microbiota networks were more dispersed than the EF networks, suggesting that the microbiota found in the EF could be more stable (Naqvi et al. 2010). Only one previous study compared the EM of EF and EB, indicating that the microbiota composition in EF does not fully reflect that in the EB (Liu et al. 2018).

To our knowledge, this is the first observational multicentre study to prospectively analyse EM composition in two types of endometrial samples (EF and EB) obtained simultaneously from the same patient and assess the association of the uterine microbial environment with pregnancy outcomes in infertile patients undergoing IVF. The data presented here are robust because both sample types were sequenced from the same bacterial 16S rRNA hypervariable regions and analysed using the same bioinformatics pipeline. However, the high sensitivity of this technology might detect DNA contamination, which can confound the interpretation of microbiome data (Eisenhofer et al. 2019), specifically in low-biomass sites such as the uterus. Therefore, we ensured that contaminating reads, but not sample-related reads, were removed from downstream analyses. For this purpose, we first classified samples as detectable or non-detectable, excluding samples that clustered with and had a roughly equivalent amplicon concentration to the blanks. Another strength of our work is that endometrial receptivity was analysed, and personalised embryo transfer was performed to synchronise endometrial receptivity with embryo development, avoiding displacement of the implantation window. Additionally, all samples were collected in a hormone replacement therapy cycle, preventing the potential bias introduced by different hormonal status. Hormonal therapy may have influenced the endometrial microbiome, but the hormonal regimen and sampling day were consistent between the sample collection analysis and embryo transfer cycles.

The main limitation of our work was the small number of BP and CM outcomes. Moreover, transcervical collection of the samples may have influenced the endometrial microbial results. Caution was taken to avoid contact of the catheter with the vaginal walls during sample collection, and although cervical contamination cannot be fully discarded, there is no other way to clinically access the endometrium. Moreover, it is now accepted that the female reproductive tract presents a continuum of microbiota (C. Chen et al. 2017), so even if some contamination from the cervix was carried over into the endometrial samples, the resulting microbial profiles would be consistent with the microbial environment of the uterine cavity. Another factor to be considered is that while sample collection and microbial analysis were performed in a cycle before embryo transfer, the microbiome may change over time.

## CONCLUSIONS

We conclude that the presence of pathogenic bacteria such as *Atopobium, Bifidobacterium, Chryseobacterium, Gardnerella, Haemophilus, Klebsiella, Neisseria, Staphylococcus*, and *Streptococcus* in the endometrium together with depletion of *Lactobacillus* spp. is associated with impaired reproductive function. These data indicate that the EM should be considered as a possible emerging cause of implantation failure and/or pregnancy loss. More studies are needed to analyse the mechanism of how pathogenic bacteria might affect embryo implantation.

## Data Availability

All raw data from this study is deposited at the Sequence Read Archive (accession code PRJNA691300) and will be publicly available upon acceptance of the manuscript.

## DECLARATIONS

### Ethics approval and consent to participate

Ethical approval was given by the corresponding local Ethics Committees to the protocol with reference IGX1-MIC-CS-17-05 as follows: Western Institutional Review Board (IVF-Florida, USA, study code 1176556, July 17, 2017; RMA Connecticut, USA, study code 1177331, July 31, 2017; Dominion Fertility, USA, study code 1176555, July 22, 2017; Missouri Center for Reproductive Medicine, USA, study code 1179405, October 10, 2017; Pacific Centre for Reproductive Medicine, Canada, study code 1179667, December 13, 2017); Comite de Etica de la Investigación con Medicamentos del Hospital Universitario de la Princesa (ProcreaTec, Spain, register number 3133, July 13, 2017); Comite de Etica de la Investigacion Costa del Sol (Clinica Fertia, Spain, code 006_jun17_PI-ERA-Microbioma, June 29, 2017); Comite de Bioetica del Instituto de Investigaciones Clinicas Rosario (Gestanza Medicina Reproductiva, Argentina, March 27, 2018); Comite de Etica en Investigación Centro de Educacion Medica e Investigaciones Clinicas “Norberto Quirno” (Pregna Medicina Reproductiva, Argentina, October 13, 2017); Human Medical Research Ethics Committee (HMREC) for Private Healthcare and Research Centres Malaysia (Alpha IVF & Women’s Specialists Centre, Malaysia, October 3, 2017); Oak Clinic Group’s Ethics Committee (Oak Clinic Sumiyoshi, Japan, May 29, 2017); Comite de Etica en Investigación New Hope Fertility Center (New Hope Fertility Center, Mexico, Register number RA-2017-04, June 9, 2017); Uskudar Universitesi Girisimsel Olamayan Arastirmalar Etik Kurulum (Bahceci Group, Turkey, January 25, 2018). All participants provided written informed consent. Data were monitored by clinical research associates through on-site and/or remote visits, and the database was properly curated by the data manager. All queries were duly solved and closed.

## Consent for publication

Not applicable

## Availability of data and materials

All raw data from this study can be found at the Sequence Read Archive (accession code PRJNA691300).

## Acknowledgements

We thank Sheila M. Cherry, PhD, ELS, President and Senior Editor from Fresh Eyes Editing LLC, for her excellent work editing this manuscript.

## Author contributions

IM and CS contributed to the conception, design, analysis and interpretation of the data. IGG and DPV contributed to the acquisition, analysis and interpretation of the data. MGM participated in the acquisition and analysis of the data. MB, MJB, ST, EP, MD, MWL, GM, MA, ML, AI, MPO, AC, and KS contributed to sample collection and acquisition of data. DB contributed to acquisition, analysis and interpretation of the data. CG contributed to design and acquisition of data. DV contributed to the design and data analysis. FV contributed to the conception and interpretation of the data. IM, IGG, and CS drafted the work. All authors have substantively revised the manuscript and approved the submitted version.

## SUPPLEMENTARY FIGURES AND TABLES

**Supplementary Figure 1:** Taxa found in endometrial fluid (A) and endometrial biopsy (B) samples.

**Supplementary Figure 2:** Taxa enriched in patients with different reproductive outcomes.

**Supplementary Table 1:** Sociodemographic and clinical characteristics of study participants.

**Supplementary Table 2:** Clinical variables in patients with different reproductive outcomes

**Supplementary Table 3:** Sequencing reads obtained from endometrial fluid and endometrial biopsy samples.

**Supplementary Table 4:** Bacteria detected in endometrial fluid and endometrial biopsy samples after filtering for low abundance taxa and potential contaminants.

