## Supplementary table 1 for "Endometrial microbiota composition is associated with reproductive outcome in infertile patients"

**Supplementary Table 1.** Sociodemographic and clinical characteristics of study participants.

|  | Total  (n=342) | Live Birth  (n=141) | No Pregnancy  (n=144) | Clinical Miscarriage  (n=28) | Biochemical Pregnancy (n=27) |
| --- | --- | --- | --- | --- | --- |
| Age (y) | 36.01 ± 4.98 | 35.28 ± 4.77 | 36.63 ± 5.08 | 35.96 ± 5.17 | 36.89 ± 5.05 |
| Body-mass index (kg/m^2^) | 23.26 ± 2.97 | 23.28 ± 2.98 | 22.93 ± 2.94 | 24.39 ± 3.16 | 23.96 ± 2.54 |
| Ethnicity |  |  |  |  |  |
| African | 9 (2.63) | 2 (1.42) | 5 (3.47) | 0 (0) | 2 (7.41) |
| Ashkenazi | 3 (0.88) | 1 (0.71) | 1 (0.69) | 0 (0) | 1 (3.7) |
| Caucasian | 196 (57.31) | 87 (61.7) | 77 (53.47) | 16 (57.14) | 15 (55.56) |
| East Asian | 48 (14.04) | 13 (9.22) | 26 (18.06) | 3 (10.71) | 5(18.52) |
| Hispanic | 39 (11.4) | 14 (9.93) | 18 (12.5) | 6 (21.43) | 1 (3.7) |
| Mulatto | 2 (0.58) | 1 (0.71) | 1 (0.69) | 0 (0) | 0 (0) |
| South Asian | 8 (2.34) | 5 (3.55) | 1 (0.69) | 1 (3.57) | 1 (3.7) |
| Other | 37 (10.82) | 18 (12.77) | 15 (10.42) | 2 (7.14) | 2 (7.14) |
| No. previous pregnancies | 0.96 ± 1.41 | 0.98 ± 1.36 | 0.87 ± 1.3 | 1.14 ± 2.17 | 1.07 ± 1.27 |
| No. previous term pregnancies | 0.2 ± 0.45 | 0.23 ± 0.5 | 0.16 ± 0.37 | 0.14 ± 0.59 | 0.26 ± 0.45 |
| No. previous miscarriages | 0.43 ± 1 | 0.43 ± 0.97 | 0.38 ± 1.02 | 0.61 ± 1.2 | 0.48 ± 0.94 |
| No. previous voluntary termination of pregnancies | 0.06 ± 0.29 | 0.06 ± 0.26 | 0.08 ± 0.34 | 0.04 ± 0.19 | 0 ± 0 |
| No. previous ectopic pregnancies | 0.09 ± 0.34 | 0.08 ± 0.32 | 0.1 ± 0.38 | 0.07 ± 0.26 | 0.07 ± 0.27 |
| No. previous stillbirths | 0 ± 0.05 | 0 ± 0 | 0.01 ± 0.08 | 0 ± 0 | 0 ± 0 |
| No. previous live births | 0.20 ± 0.46 | 0.24 ± 0.51 | 0.16 ± 0.37 | 0.14 ± 0.59 | 0.3 ± 0.54 |
| No. implantation failures | 1.99 ± 1.70 | 1.87 ± 1.68 | 2.24 ± 1.75 | 1.57 ± 1.53 | 1.63 ± 1.67 |
| ART indication |  |  |  |  |  |
| Advanced maternal age | 38 (11.11) | 13 (9.22) | 14 (97.22) | 3 (10.71) | 6 (22.22) |
| Male factor | 30 (8.77) | 18 (12.76) | 9 (6.25) | 2 (7.14) | 1 (3.7) |
| RPL | 3 (0.88) | 2 (1.42) | 1 (0.69) | 0 (0) | 0 (0) |
| RIF | 11 (3.22) | 4 (2.84) | 5 (3.47) | 1 (3.57) | 0 (0) |
| Tubal factor | 9 (2.63) | 3 (2.13) | 3 (2.08) | 2 (7.14) | 1 (3.7) |
| Uterine factor | 1 (0.29) | 0 (0) | 0 (0) | 0 (0) | 1 (3.7) |
| Ovarian pathology | 20 (5.85) | 11 (7.8) | 8 (5.55) | 1 (3.57) | 0 (0) |
| Endometriosis | 7 (2.05) | 4 (2.84) | 3(2.08) | 0 (0) | 0 (0) |
| Genetic factor | 8 (2.34) | 2 (1.42) | 4 (2.77) | 1 (3.57) | 1 (3.7) |
| Unexplained infertility | 28 (8.19) | 13 (9.22) | 10 (6.94) | 3 (10.71) | 1 (3.7) |
| No indication | 1 (0.29) | 1 (0.7) | 0 (0) | 0 (0) | 0 (0) |
| > 1 indication | 186 (54.38) | 70 (49.64) | 87 (60.42) | 15 (53.57) | 16 (59.26) |
| FSH (IU/mL) | 7.59 ± 3.85 | 7.18 ± 3.53 | 8.14 ± 4.71 | 7 ± 1.95 | 7.87 ± 2.77 |
| AMH (ng/mL) | 2.98 ± 3.29 | 2.82 ± 3.05 | 2.89 ± 3.48 | 3.89 ± 5.39 | 3.06 ± 1.5 |
| Sperm concentration (millions/mL) | 53.03 ± 51 | 49.88±44.51 | 54.98±49.62 | 41.76±31.99 | 63.97±77.66 |

Data are expressed as mean ± SD or n (%).

Analysis of variance was used to compare numerical variables in the four groups, whereas the independent samples Student's t-test was used to compare quantity variables between groups. Chi-Square and Fisher’s exact tests were used to compare categorical variables. Bonferroni was used for multiple comparison testing.

AMH, anti-Müllerian hormone; ART, assisted reproductive treatment; FSH, follicle stimulating hormone; RIF, recurrent implantation failure; RPL, recurrent pregnancy loss.
