## Supplementary table 2 for "Endometrial microbiota composition is associated with reproductive outcome in infertile patients"

**Supplementary Table 2. Clinical variables in patients with different reproductive outcomes.**

|  | Live Birth  (n=141) | | No Pregnancy  (n=144) | Clinical Miscarriage  (n=28) | Biochemical Pregnancy (n=27) |
| --- | --- | --- | --- | --- | --- |
| **ART information** |  | |  |  |  |
| Treatment |  | |  |  |  |
| IVF | 17 (12.06) | | 16 (11.11) | 3 (10.71) | 0 (0) |
| IVF/ICSI | 124 (87.94) | | 128 (88.89) | 25 (89.29) | 27 (100) |
| Stimulation protocol |  | |  |  |  |
| Long | 40 (28.37) | | 27 (18.75) | 6 (21.43) | 5 (18.51) |
| Short | 85 (60.28) | | 84 (58.33) | 18 (64.28) | 15 (55.55) |
| Unknown | 16 (11.35) | | 33 (22.92) | 4 (14.29) | 7 (25.93) |
| **COS cycle** |  | |  |  |  |
| Egg source |  | |  |  |  |
| Donated | 26 (18.44) | | 37 (25.69) | 8 (28.57) | 11 (40.74) |
| Own | 115 (81.56) | | 107 (74.31) | 20 (71.43) | 16 (59.26) |
| Ovulation induction | |  |  |  |  |
| Agonist | 41 (29.07) | | 28 (19.44) | 7 (25) | 5 (18.51) |
| Double triggering | 19 (13.47) | | 11 (7.63) | 4 (14.28) | 2 (7.40) |
| hCG | 61 (43.26) | | 71 (49.31) | 13 (46.43) | 10 (37.03) |
| Unknown | 20 (14.18) | | 34 (23.61) | 4 (14.29) | 10 (37.04) |
| No. antral follicles | 17.36 ± 8.95 | | 14.75 ± 6.65 | 18.17 ± 11.57 | 17.78 ± 10.25 |
| No. metaphase II oocytes | 12.31 ± 6.96 | | 10.43 ± 5.96 | 12.72 ± 8.04 | 11.88 ± 7.42 |
| **ERA cycle** |  | |  |  |  |
| Endogenous P4 value nm/mL | 0.34 ± 0.26 | | 0.37 ± 0.27 | 0.38 ± 0.36 | 0.34 ± 0.15 |
| E2 days in ERA cycle | 17.89 ± 4.33 | | 18.15 ± 4.93 | 17.74 ± 4.61 | 18.89 ± 5.36 |
| Total hours P4 in ERA cycle | 122.01 ± 7.46 | | 120.7 ± 5.76 | 123.47 ± 6.9 | 121.85 ± 3.33 |
| Progesterone hours recommended for ET | 129.38 ± 11.95 | | 127.3 ± 11.56 | 131.64 ± 11.75 | 130.59 ± 13.41 |
| **ET cycle** |  | |  |  |  |
| Antibiotic intake in ET | 22 (15.60) | | 18 (12.5) | 4 (14.29) | 3 (11.11) |
| Probiotics before ET | 6 (4.26) | | 12 (8.33) | 2 (7.14) | 4 (14.81) |
| Total hours progesterone in ET cycle | 129.38 ± 12.01 | | 127.22 ± 11.02 | 131.8 ± 11.97 | 131.29 ± 13.63 |
| E2 days in ET cycle | 19.22 ± 5.53 | | 19.69 ± 5.87 | 17.39 ± 6.68 | 21.59 ± 5.91 |
| Endogenous P4 value nm/mL | 0.33 ± 0.23 | | 0.36 ± 0.24 | 0.33 ± 0.19 | 0.32 ± 0.12 |
| Days between sample collection and ET | 84.34 ± 73.43 | | 95.26 ± 87.81 | 77.82 ± 34.69 | 105.81 ± 98.7 |
| Embryo day at ET |  | |  |  |  |
| 5 | 115 (81.56) | | 120 (83.33) | 23 (82.14) | 20 (74.07) |
| 6 | 26 (18.44) | | 24 (16.67) | 5 (17.86) | 7 (25.93) |
| No. embryos transferred | 1.18 ± 0.38 | | 1.12 ± 0.33 | 1.14 ± 0.36 | 1.15 ± 0.36 |
| Embryo stage |  | |  |  |  |
| Cavitated blastocyst | 12 (8.51) | | 14 (9.72) | 3 (10.71) | 5 (18.52) |
| Early blastocyst | 1 (0.71) | | 6 (4.17) | 1 (3.57) | 1 (3.7) |
| Expanded blastocyst | 57 (40.43) | | 54 (37.5) | 13 (46.43) | 13 (48.15) |
| Hatched blastocyst | 11 (7.8) | | 4 (2.78) | 0 (0) | 3 (11.11) |
| Hatching blastocyst | 60 (42.55) | | 65 (45.14) | 11 (39.29) | 5 (18.52) |
| Unknown | 0 (0) | | 1 (0.69) | 0 (0) | 0 (0) |
| Embryo quality (inner cell mass/trophectoderm) |  | |  |  |  |
| AA | 24 (17.02) | | 29 (20.14) | 5 (17.86) | 3 (11.11) |
| AB | 20 (14.18) | | 23 (15.97) | 5 (17.86) | 3 (11.11) |
| AC | 2 (1.42) | | 0 (0) | 0 (0) | 0 (0) |
| BA | 8 (5.67) | | 11 (7.64) | 3 (10.71) | 3 (11.11) |
| BB | 48 (34.04) | | 38 (26.39) | 9 (32.14) | 10 (37.04) |
| BC | 22 (15.6) | | 21 (14.58) | 4 (14.29) | 2 (7.41) |
| BD | 0 (0) | | 0 (0) | 0 (0) | 1 (3.7) |
| CA | 2 (1.42) | | 1 (0.69) | 1 (3.57) | 0 (0) |
| CB | 5 (3.55) | | 3 (2.08) | 0 (0) | 0 (0) |
| CC | 8 (5.67) | | 12 (8.33) | 1 (3.57) | 3 (11.11) |
| Unknown | 2 (1.41) | | 6 (4.16) | 0 (0) | 2 (7.4) |

Data are expressed as mean ± SD or n (%).

Analysis of variance was used to compare numerical variables in the four groups, whereas an independent samples Student's t-test was used to compare quantity variables between groups. Chi-Square and Fisher’s Exact tests were used to compare categorical variables. Bonferroni was used for the multiple comparison test.

ART, assisted reproductive treatment; ERA, endometrial receptivity analysis; COS, controlled ovarian stimulation; ET, embryo transfer; ICSI, intracytoplasmic sperm injection; IVF, in vitro fertilisation; P4, progesterone; hCG, human chorionic gonadotropin; E2, estradiol.
