## Supplementary table 3 for "Endometrial microbiota composition is associated with reproductive outcome in infertile patients"

**Supplementary Table 3.** Sequencing reads obtained from endometrial fluid and endometrial biopsy samples.

| ENDOMETRIAL FLUID | | | | |
| --- | --- | --- | --- | --- |
| RUN | Total reads | Mapped reads | Filtered reads | OTUs No. |
| Auto_user_S5XL-IGENOMIX-681-RUN1-niERA-MB_060319_Ion_16S_Metagenomics_IonXpress_1323_1408-10.100.40.97 | 234769.05 | 87643.64 | 72768.52 | 132.33 |
| Auto_user_S5XL-00245-780-RUN2-niERA-MB_070319_Ion_16S_Metagenomics_IonXpress_1703_1632-10.100.40.146 | 320129.12 | 142889.12 | 121855.17 | 168.71 |
| Auto_user_S5XL-00245-784-RUN3-niERA-MB_080319_Ion_16S_Metagenomics_IonXpress_1707_1640-10.100.40.146 | 331989.15 | 123320.72 | 105983.3 | 175.65 |
| Auto_user_S5XL-IGENOMIX-691-RUN4-niERA-MB_120319_Ion_16S_Metagenomics_IonXpress_1336_1428-10.100.40.97 | 331209.44 | 124661.54 | 101033.59 | 171.95 |
| Auto_user_S5XL-IGENOMIX-695-RUN5-niERA-MB_130319_Ion_16S_Metagenomics_IonXpress_1339_1436-10.100.40.97 | 387134.78 | 179377.75 | 137956.98 | 159.98 |
| Auto_user_S5XL-00245-798-RUN6-niERA-MB_140319_Ion_16S_Metagenomics_IonXpress_1725_1668-10.100.40.146 | 378634.54 | 147026.34 | 102549.37 | 179.95 |
| Auto_user_S5XL-IGENOMIX-706-RUN7-niERA-MB_210319_Ion_16S_Metagenomics_IonXpress_1356_1458-10.100.40.97 | 394659.98 | 151487.98 | 115704.1 | 169.66 |
| Auto_user_S5XL-IGENOMIX-715-RUN8-niERA-MB_270319_Ion_16S_Metagenomics_IonXpress_1372_1476-10.100.40.97 | 325167.75 | 137347.73 | 94428.65 | 176.52 |
| Auto_user_S5XL-IGENOMIX-885-RUN9_2-niERA-MB_270719_Ion_16S_Metagenomics_IonXpress_1690_1823-10.100.40.97 | 387438.04 | 173307.93 | 89311.42 | 72.98 |
| Auto_user_S5XL-00245-891-RUN10-niERA-MB_260719_Ion_16S_Metagenomics_IonXpress_1947_1864-10.100.40.146 | 376633.96 | 157690.93 | 115462.98 | 92.44 |
| Auto_user_S5XL-00245-942-Run11_niERA-MB_Ion_16S_Metagenomics_IonXpress_2028_1967-10.100.40.146 | 346387.93 | 152794.84 | 107344.71 | 95.73 |
| Auto_user_S5XL-00245-966-RUN12-niERA-MB_211019_Ion_16S_Metagenomics_IonXpress_2055_2019-10.100.40.146 | 366068.87 | 162291.44 | 102723.64 | 84.4 |
| Auto_user_S5XL-00245-986-RUN13-niERA-MB_05112019_Ion_16S_Metagenomics_IonXpress_2078_2059-10.100.40.146 | 347317.8 | 144210.83 | 96974.37 | 84.94 |
| Auto_user_S5XL-IGENOMIX-110-Run_9_Ion_16S_Metagenomics_IonXpress_226_226-10.100.40.97 | 302134.18 | 110782.43 | 96429.35 | 55.22 |
| Auto_user_S5XL-IGENOMIX-155-RUN11_2_09112017_Ion_16S_Metagenomics_IonXpress_337_316-10.100.40.97 | 110050.43 | 40634.72 | 27960.7 | 46.32 |
| Auto_user_S5XL-IGENOMIX-177-MICROBIOMA_12_Ion_16S_Metagenomics_IonXpress_376_360-10.100.40.97 | 138513.16 | 36893.89 | 28045.57 | 25.14 |
| Auto_user_S5XL-IGENOMIX-355-RUN18_2_Microbiome_Ion_16S_Metagenomics_IonXpress_646_730-10.100.40.97 | 187788.05 | 61802.79 | 44261.8 | 113.39 |
| Auto_user_S5XL-IGENOMIX-610-RUN24_MICROBIOMA_Ion_16S_Metagenomics_IonXpress_1207_1264-10.100.40.97 | 175361.38 | 66923.75 | 57109.18 | 120.04 |
| MEAN | 302299.31 | 122282.69 | 89883.52 | 118.07 |
| MAX | 394659.98 | 179377.75 | 137956.98 | 179.95 |
| MIN | 110050.43 | 36893.89 | 27960.70 | 25.14 |
| ENDOMETRIAL BIOPSY | | | | |
| RUN | Total reads | Mapped reads | Filtered reads | OTUs No. |
| Auto_user_GSS5PR-0157-130-ESEMMA20190924-1-1_294_264-10.100.40.148 | 374648.54 | 139257.49 | 100325.77 | 30.83 |
| Auto_user_S5XL-00245-940-ESEMMA20191001-1-1_2026_1963-10.100.40.146 | 334764.84 | 116638.03 | 92541.74 | 27.03 |
| Auto_user_S5XL-00245-955-ESEMMA20191014-1-1_2043_1994-10.100.40.146 | 237889.03 | 77143.00 | 61650.59 | 34.46 |
| Auto_user_S5XL-00245-982-ESEMMA20191030-1-1_2074_2051-10.100.40.146 | 283484.28 | 113645.53 | 77784.31 | 30.11 |
| Auto_user_S5XL-IGENOMIX-1057-ESEMMA20191108-1-1_1932_2168-10.100.40.97 | 298889.82 | 122723.1 | 92725.08 | 35.28 |
| Auto_user_S5XL-00245-1080-ESEMMA20200116-1-1_2194_2251-10.100.40.146 | 383769.88 | 188452.55 | 162653.42 | 30.85 |
| Auto_user_S5XL-IGENOMIX-1162-ESEMMA20200122-1-1_2084_2381-10.100.40.97 | 430675.21 | 159725.59 | 142480.24 | 25.15 |
| Auto_user_S5XL-IGENOMIX-1173-ESEMMA20200128-1-1_2099_2403-10.100.40.97 | 350212.78 | 139095.49 | 117724.95 | 30.27 |
| Auto_user_S5XL-IGENOMIX-1182-ESEMMA20200204-1-1_2115_2421-10.100.40.97 | 275900.98 | 108071.28 | 95751.79 | 27.04 |
| Auto_user_S5XL-00245-1138-ESEMMA20200302-1-1_2270_2376-10.100.40.146 | 386357.05 | 104739.46 | 91759.44 | 22.22 |
| MEAN | 335659.24 | 126949.15 | 103539.73 | 29.32 |
| MAX | 430675.21 | 188452.55 | 162653.42 | 35.28 |
| MIN | 237889.03 | 77143.00 | 61650.59 | 22.22 |
