## Supplementary table 4 for "Endometrial microbiota composition is associated with reproductive outcome in infertile patients"

**Supplementary Table 4.** Bacteria detected in endometrial fluid and endometrial biopsy samples after filtering for low abundance taxa and potential contaminants.

| ENDOMETRIAL FLUID | Sample  % | Sample % higher than 1% | Sample % higher than 0.1% |
| --- | --- | --- | --- |
| **k__Bacteria;p__Firmicutes;c__Bacilli;o__Lactobacillales;f__Lactobacillaceae;g__Lactobacillus** | 100.00% | 94.82% | 100.00% |
| k__Bacteria;p__Actinobacteria;c__Actinobacteria;o__Actinomycetales;f__Streptomycetaceae;g__Streptomyces | 95.85% | 68.39% | 87.05% |
| **k__Bacteria;p__Firmicutes;c__Bacilli;o__Lactobacillales;f__Streptococcaceae;g__Streptococcus** | 98.96% | 20.73% | 59.07% |
| **k__Bacteria;p__Actinobacteria;c__Actinobacteria;o__Bifidobacteriales;f__Bifidobacteriaceae;g__Gardnerella** | 96.37% | 19.69% | 39.38% |
| **k__Bacteria;p__Actinobacteria;c__Actinobacteria;o__Bifidobacteriales;f__Bifidobacteriaceae;g__Bifidobacterium** | 73.06% | 15.03% | 22.28% |
| **k__Bacteria;p__Actinobacteria;c__Actinobacteria;o__Actinomycetales;f__Corynebacteriaceae;g__Corynebacterium** | 98.45% | 13.47% | 56.99% |
| **k__Bacteria;p__Firmicutes;c__Bacilli;o__Bacillales;f__Staphylococcaceae;g__Staphylococcus** | 94.30% | 12.95% | 38.86% |
| **k__Bacteria;p__Bacteroidetes;c__Bacteroidia;o__Bacteroidales;f__Prevotellaceae;g__Prevotella** | 83.42% | 8.81% | 23.83% |
| k__Bacteria;p__Bacteroidetes;c__Flavobacteriia;o__Flavobacteriales;f__[Weeksellaceae];g__Chryseobacterium | 76.68% | 8.29% | 25.91% |
| **k__Bacteria;p__Actinobacteria;c__Coriobacteriia;o__Coriobacteriales;f__Coriobacteriaceae;g__Atopobium** | 55.96% | 8.29% | 10.88% |
| k__Bacteria;p__Firmicutes;c__Clostridia;o__Clostridiales;f__Clostridiaceae;g__Clostridium | 68.91% | 5.70% | 15.03% |
| **k__Bacteria;p__Actinobacteria;c__Actinobacteria;o__Actinomycetales;f__Propionibacteriaceae;g__Propionibacterium** | 97.93% | 4.66% | 46.11% |
| **k__Bacteria;p__Firmicutes;c__Clostridia;o__Clostridiales;f__[Tissierellaceae];g__Anaerococcus** | 65.80% | 3.11% | 15.03% |
| **k__Bacteria;p__Proteobacteria;c__Gammaproteobacteria;o__Pasteurellales;f__Pasteurellaceae;g__Haemophilus** | 66.84% | 2.07% | 15.03% |
| **k__Bacteria;p__Actinobacteria;c__Actinobacteria;o__Actinomycetales;f__Microbacteriaceae;g__Microbacterium** | 83.94% | 1.04% | 22.28% |
| ENDOMETRIAL BIOPSY | Sample % | Sample % higher than 1% | Sample % higher than 0.1% |
| **k__Bacteria;p__Firmicutes;c__Bacilli;o__Lactobacillales;f__Lactobacillaceae;g__Lactobacillus** | 100.00% | 89.95% | 99.47% |
| **k__Bacteria;p__Actinobacteria;c__Actinobacteria;o__Actinomycetales;f__Propionibacteriaceae;g__Propionibacterium** | 100.00% | 36.51% | 82.54% |
| **k__Bacteria;p__Firmicutes;c__Bacilli;o__Lactobacillales;f__Streptococcaceae;g__Streptococcus** | 94.18% | 26.98% | 60.85% |
| **k__Bacteria;p__Actinobacteria;c__Actinobacteria;o__Actinomycetales;f__Corynebacteriaceae;g__Corynebacterium** | 95.24% | 25.93% | 72.49% |
| **k__Bacteria;p__Actinobacteria;c__Actinobacteria;o__Actinomycetales;f__Microbacteriaceae;g__Microbacterium** | 57.14% | 24.34% | 48.68% |
| k__Bacteria;p__Proteobacteria;c__Betaproteobacteria;o__Burkholderiales;f__Oxalobacteraceae;g__Cupriavidus | 60.32% | 23.81% | 48.15% |
| **k__Bacteria;p__Actinobacteria;c__Actinobacteria;o__Bifidobacteriales;f__Bifidobacteriaceae;g__Gardnerella** | 92.59% | 20.63% | 43.92% |
| **k__Bacteria;p__Firmicutes;c__Bacilli;o__Bacillales;f__Staphylococcaceae;g__Staphylococcus** | 94.18% | 19.05% | 58.73% |
| **k__Bacteria;p__Actinobacteria;c__Actinobacteria;o__Bifidobacteriales;f__Bifidobacteriaceae;g__Bifidobacterium** | 58.73% | 15.87% | 31.75% |
| **k__Bacteria;p__Bacteroidetes;c__Bacteroidia;o__Bacteroidales;f__Prevotellaceae;g__Prevotella** | 55.56% | 10.05% | 30.16% |
| **k__Bacteria;p__Actinobacteria;c__Coriobacteriia;o__Coriobacteriales;f__Coriobacteriaceae;g__Atopobium** | 49.74% | 8.47% | 16.93% |
| k__Bacteria;p__Proteobacteria;c__Gammaproteobacteria;o__Enterobacteriales;f__Enterobacteriaceae;g__Escherichia | 87.30% | 7.41% | 33.86% |
| k__Bacteria;p__Proteobacteria;c__Gammaproteobacteria;o__Enterobacteriales;f__Enterobacteriaceae;g__Klebsiella | 29.63% | 5.29% | 7.41% |
| **k__Bacteria;p__Firmicutes;c__Clostridia;o__Clostridiales;f__[Tissierellaceae];g__Anaerococcus** | 49.21% | 3.17% | 22.75% |
| k__Bacteria;p__Firmicutes;c__Bacilli;o__Bacillales;f__Bacillaceae;g__Bacillus | 40.74% | 3.17% | 22.22% |
| k__Bacteria;p__Firmicutes;c__Clostridia;o__Clostridiales;f__[Tissierellaceae];g__Finegoldia | 39.15% | 3.17% | 20.11% |
| **k__Bacteria;p__Proteobacteria;c__Gammaproteobacteria;o__Pasteurellales;f__Pasteurellaceae;g__Haemophilus** | 38.62% | 2.65% | 21.69% |
| k__Bacteria;p__Actinobacteria;c__Actinobacteria;o__Actinomycetales;f__Micrococcaceae;g__Micrococcus | 37.04% | 2.65% | 17.46% |
| k__Bacteria;p__Proteobacteria;c__Betaproteobacteria;o__Burkholderiales;f__Comamonadaceae;g__Tepidimonas | 61.38% | 0.53% | 18.52% |
